# Genetically informed drug target prioritisation and repurposing for major depressive disorder

**DOI:** 10.1101/2025.07.15.25331558

**Authors:** Abigail R. ter Kuile, Chris Finan, Sandesh Chopade, Marion Van Vugt, Nikita Hukerikar, Serena Barral, Argyris Stringaris, Amand F. Schmidt, Karoline Kuchenbaecker, Jean-Baptiste Pingault

## Abstract

Major depressive disorder (MDD) treatments have limited efficacy and target few mechanisms, highlighting the need for innovative drug discovery. Drugs targeting genetically supported proteins are 2.6 times more likely to succeed in drug development. Here, we use genetic methods to identify and prioritise MDD drug targets, leveraging genome-wide association study (GWAS) summary statistics from >525,000 MDD cases. We derived exposure data from 10 GWAS measuring protein quantitative trait loci (pQTLs) and gene expression levels (eQTLs) in blood, cerebrospinal fluid, and brain tissues. We performed *cis*- Mendelian randomisation (MR) on 3,469 druggable targets (genes encoding proteins targeted by existing compounds or experimentally predicted to be druggable). To strengthen causal inference, we implemented robust MR estimators, colocalisation, external replication, and assessed directional consistency across tissues. We integrated MR effect directions with drug mechanisms and clinical annotations to infer potential therapeutic effects. Validation analyses showed that 82% of drugs approved for depression/anxiety had ≥1 significant MR target, compared to 51% for compounds in clinical trials. For repurposing, we prioritised 54 targets of compounds developed for other conditions with estimated beneficial effects on MDD (e.g., an inhibitor for a risk-increasing target). Ten high-priority targets of brain-penetrating compounds included ACE and NISCH (cardiovascular drugs), NDUFA2, NDUFB6, and NDUFS1 (metformin), CDK4, NTRK3, and MET (oncology inhibitors), and GLS and NOS2 (enzyme inhibitors). We found genetic evidence for established and novel MDD targets across the drug development pipeline. Novel targets point to mechanisms beyond monoaminergic systems, most with approved drugs for other conditions, offering immediate repurposing opportunities.

## Introduction

Major depressive disorder (MDD) is common and a leading cause of years lived with disability globally [1]. Current pharmacological treatments are ineffective for many with MDD, with only one-third of patients reaching remission after first-line antidepressant treatment [2]. Furthermore, 28-40% of patients are classified as having treatment-resistant depression, defined as not remitting after the prescription of two or more antidepressants [2,3]. Currently approved drugs target a narrow range of therapeutic mechanisms, with most antidepressants acting on monoaminergic neurotransmitter systems [4]. The lack of novel pharmacological treatments for MDD highlights the pressing need for innovative strategies to facilitate efficient drug discovery.

Human genetics offers a promising avenue to guide the development of pharmacological treatments. Drugs acting on targets with supporting human genetic evidence are 2.6 times more likely to succeed in drug discovery pipelines and progress into clinical practice than those without [5–7]. This is reflected in recent FDA approvals: 63% of drugs approved in the past decade had genetic support [8]. Given that genes encode proteins, which constitute over 90% of drug targets, genetic methods can identify disease-associated protein targets suitable for repurposing existing drugs or for novel development opportunities [9,10]. To accelerate discovery and repurposing, an efficient strategy involves focusing on the ‘druggable genome’ rather than all protein-coding genes, reducing the multiple testing burden [9–11]. The druggable genome comprises genes encoding proteins targeted by drugs that are already approved, those at the clinical stage of drug development, or those potentially druggable based on experimental evidence [9].

Mendelian randomisation (MR) enables scanning of the druggable genome by using genetic variants as instruments to estimate the effect of altering each drug target’s activity on disease risk [12]. For each target protein, genetic variants selected as instruments lie within and around the gene encoding that protein (*cis*-region) [11]. These instruments are associated with individual variation in a molecular trait in a given tissue, including changes in protein values or mRNA expression [10]. *Cis*-MR leverages data from genome-wide association studies (GWAS) to obtain the effects of each selected variant on both the exposure (target protein activity) and the outcome (MDD). To infer exposure-outcome causal effects, MR assumes that variant effects on the outcome operate through the exposure and are independent of confounders [12]. Under these assumptions, a significant MR estimate suggests that modulating the protein corresponding to the instrumented *cis*-region may influence the disease process, making it a candidate drug target for that outcome [10,12]. MR has key advantages for identifying drug targets. First, the random assortment of genetic variants at conception mimics treatment allocation in randomised controlled trials, strengthening causal inference regarding target effects on disease outcomes [13]. Second, MR provides the direction of effect to infer whether activating or inhibiting a target would mitigate disease risk [14]. The effect direction can then be combined with pharmacological data on existing drugs to identify target-compound pairs with aligned mechanisms, thus directly aiding repurposing [14,15].

To conduct drug target MR, a large-scale GWAS of the disease outcome of interest is fundamental. The latest Psychiatric Genomics Consortium (PGC) Phase 3 MDD GWAS offers unprecedented opportunities in this regard. The meta-analysis found 635 genomic loci associated with MDD, of which 293 are novel [16]. Alongside identifying over 300 high-confidence genes and associated pathways, the study performed drug target enrichment analyses [16]. Using sets of genes encoding drug targets and grouping them by drug classes showed that antidepressant and antipsychotic classes were enriched for MDD-associated variants [16]. Additionally, 18 individual compounds approved mainly for indications other than depression were enriched, including antipsychotics, treatment for cancer, anxiety (pregabalin), and narcolepsy (modafinil) [16]. However, as noted by the authors, enrichment analysis is not directional and cannot distinguish whether implicated compounds might increase or decrease MDD symptoms [16]. Enrichment of established antidepressant drug class alongside potential repurposing opportunities underscores the value of this large-scale GWAS dataset for a detailed and directional drug target MR interrogation.

While initial druggable genome *cis*-MR applications to MDD have provided a foundation by inferring directional effects [17,18], opportunities remain to advance and strengthen this approach. The current study addresses this by introducing an integrated approach that leverages several methodological advances to robustly identify and prioritise targets. First, we enhance statistical power by using the latest large-scale PGC Phase 3 MDD GWAS [16]. Second, we expand the scope of the druggable genome beyond established targets to include novel candidates supported by experimental evidence of druggability. Third, we evaluate target effects across blood, cerebrospinal fluid (CSF), and multiple brain regions, beyond previous brain-restricted analyses [17,18]. Finally, our framework strengthens the validation of genetic evidence by employing multiple robust MR estimators, colocalisation, external replication, and tissue-specific directional consistency.

To identify a tractable set of high-priority targets from our *cis*-MR findings for experimental and clinical validation, we integrate genetic evidence with clinical and pharmacological data. Such a framework has been applied to other non-psychiatric complex diseases, evaluating genetic robustness, clinical relevance, and pharmacological properties of target-compound pairs [14,15,19]. These studies integrated target effect directions with pharmacological mechanisms and clinical profiles of existing drugs [14,15,19]. Aligning the genetically estimated benefit of modulating a target with the mechanisms of drugs approved for other indications can identify high-confidence repurposing candidates [14,15]. The approach was validated by genetically recapitulating established pharmacological treatments for diseases, known as ‘positive controls’ [14,15]. Applying this framework to MDD is particularly pertinent, where therapeutic mechanisms are thought to be complex [4]. Addressing a gap in prior MDD drug target MR investigations [17,18], we also evaluate established antidepressant targets as positive controls to benchmark analyses and strengthen prioritisation of novel candidates.

We applied an integrated genetic and pharmacological framework to systematically identify and prioritise MDD drug targets within the druggable genome. First, we aimed to identify targets using *cis*-MR and assess the genetic robustness of findings through multiple sensitivity analyses. Second, we sought to validate our approach using established MDD target-compound pairs as positive controls. Third, we aimed to characterise repurposing opportunities by integrating genetic effects with drug mechanisms of action and clinical annotations (e.g., primary indications and side effects).

## Methods

### Outcome and exposure data

#### GWAS summary statistics

For our outcome, we used summary statistics from the PGC MDD phase 3 GWAS meta-analysis (525,197 cases and 3,362,335 controls) of European genetic ancestry (PGC MDD-3) [16]. We derived exposure genetic instruments from 10 GWAS of protein or transcript values measured in blood [20–22], cerebrospinal fluid (CSF) [23], or multiple brain regions [24,25] (Supplementary Table 1). These genetic variants are quantitative trait loci (QTLs) strongly associated with changes in protein levels (pQTLs) or mRNA expression (eQTLs) in each tissue [10]. To maximise protein coverage and enable cross-platform validation [26], blood plasma proteomic data was derived from two pQTL GWAS measured using different protein assay platforms: deCODE (SomaScan) and UK Biobank Pharma Proteomics Project (Olink; Supplementary Methods) [20,21]. We analysed each protein/transcript GWAS separately (hereafter referred to as a ‘panel’).

#### Drug target data

We selected 3,652 genes encoding drug target proteins for *cis*-MR analysis using hierarchical druggability assessments from Open Targets [27]. We included all targets with clinical trial evidence from ChEMBL [28], with the following hierarchy: approved drugs (phase IV, already ‘drugged’), advanced clinical development (phases II-III), and early clinical development (phase I). For targets with only experimental evidence (i.e. potentially druggable), we retained those within the top tier of experimental quality assessments from Open Targets hierarchical scoring (Supplementary Methods) to incorporate targets with promising druggability potential.

### Primary Mendelian Randomisation (MR) analysis

We used purpose-built Python packages for *cis*-MR analyses [19]. We selected genetic variants as instruments with minor allele frequencies (MAF) > 0.01, located within 200kb of the *cis*-gene [29], and with exposure GWAS p-value < 1×10^-6^. We applied MR Steiger filtering to minimise reverse causation by removing variants with a greater effect on the outcome than on the exposure. Variants were clumped (linkage disequilibrium (LD) R^2^ = 0.4), and remaining instrument correlations were accounted for by generalised least squares modelling. LD reference matrices were derived from a random subset of 5,000 UK Biobank participants of European genetic ancestry [30]. We employed the Wald ratio estimator (MR-Wald) for targets instrumented by a single genetic variant, and the generalised least squares implementation of the inverse-variance weighted estimator (MR-IVW), allowing for correlated instruments for targets with multiple variants [31]. To mitigate potential horizontal pleiotropy in our IVW analysis, we applied heterogeneity-based pruning of variants with large leverage (>3 times the mean) or outlier statistics (chi-square >11) [19,32]. We implemented fixed-effects IVW for targets with <4 IVs and random-effects for ≥4 [33,34].

A total of 3,469 targets of the 3,652 in the druggable genome had instrument(s) that passed our filtering criteria. We applied Bonferroni correction (p<3.9×10^-6^), accounting for 12,794 MR-IVW/-Wald tests across 10 panels spanning blood, brain and CSF. We used the total number of tests rather than unique targets tested, as most targets (94%) were tested more than once across panels (Supplementary Table 2).

### Sensitivity analyses and target annotation

#### MR-IVW robustness

We employed three robust methods, with different assumptions about instrument validity [35], across four models: generalised least squares implementation of MR-Egger with and without heterogeneity-based pruning [36], contamination mixture [37], and weighted median methods [38] (Supplementary Methods). We applied Bonferroni correction (p<7.2×10^-5^) for 693 significant IVW estimates from our primary analysis that we sought to test with robust MR. Robust MR estimates that were significant and directionally consistent with IVW estimates for that target provided stronger evidence for the validity of our causal estimates [35]. Targets with significant but inconsistent directional effects across IVW and robust estimators were deprioritised.

#### External pQTL replication

We performed external replication of targets significant in our primary *cis*-MR analyses in independent QTL datasets, focusing on panels with a matching molecular phenotype modality (i.e. pQTL) and tissue type (i.e., blood or brain) to our discovery panels (Supplementary Methods and Supplementary Table 3). We defined successful replication as a significant MR-IVW/-Wald effect in the replication panel that was directionally consistent with the discovery panel effect estimate. The eQTL targets were not tested for external replication as they were derived from large-scale meta-analyses for which comparably well-powered independent datasets were unavailable.

#### Colocalisation

We performed colocalisation analyses for each significant MR-IVW/-Wald target in a given QTL panel to evaluate whether molecular traits (pQTL/eQTL) and MDD share causal variants. We used the *coloc* R package and the *‘coloc.abf*’ function (Supplementary Methods) [39] with a posterior probability threshold of ≥ 0.8 to indicate strong evidence. Strong evidence for PPH4 indicates a shared causal variant between the molecular trait and MDD offering triangulation of evidence alongside *cis-*MR [29,40]. Conversely, strong PPH3 evidence indicates distinct causal variants for each trait, suggesting potential violations of MR assumptions due to, for example, linkage disequilibrium, or pleiotropic pathways through neighbouring genes [29,40].

#### Estimated therapeutic relevance

To evaluate the therapeutic relevance of drug targets with significant *cis*-MR effects, we integrated our genetic findings with drug mechanisms of action data from ChEMBL [28]. We estimated whether existing compounds targeting that protein would act in a potentially beneficial direction based on the MR-IVW/-Wald effect direction (Supplementary Methods). We classified therapeutic directional effects as 1) ‘beneficial’, where the drug’s mechanism mitigates the risk-increasing effect of the target on MDD (e.g., an inhibitor for a target whose increased expression increases MDD risk); 2) ‘adverse’, the drug’s mechanism is in the same direction as the target’s risk-increasing effect; or 3) ‘unknown’, the relationship could not be classified (e.g., for mechanisms that do not specify inhibition or activation).

#### Known depression-related drug effects

We extracted known depression-related therapeutic applications or side effects for compounds targeting significant MR-IVW/-Wald proteins. We classified drug effects using core depression terms (MDD, depression, apathy, mood, anxiety, suicidality) and broader depression-relevant terms encompassing related psychiatric and somatic symptoms (Supplementary Methods). Drug effects data were extracted from databases of drugs licensed in the UK (primary therapeutic indications, side effects, cautions, and contraindications) and ChEMBL (primary compound indications in clinical trial phases I-IV) [28]. Depression-related clinical effects, including adverse effects, suggest biological relevance. Targets can be modulated by drugs with different mechanisms (activators or inhibitors), allowing for the development of novel drugs with beneficial effects. Furthermore, as documented in databases of medications licensed in the UK, approved antidepressants have reported psychiatric side effects, including mood changes and anxiety. Thus, depression-related symptoms as side effects do not necessarily preclude therapeutic potential.

#### Target prioritisation

We evaluated significant MR-IVW/-Wald drug targets from our primary analysis by considering evidence for each target across assessments of genetic robustness and therapeutic potential (**Figure 1**).

**Figure 1:**
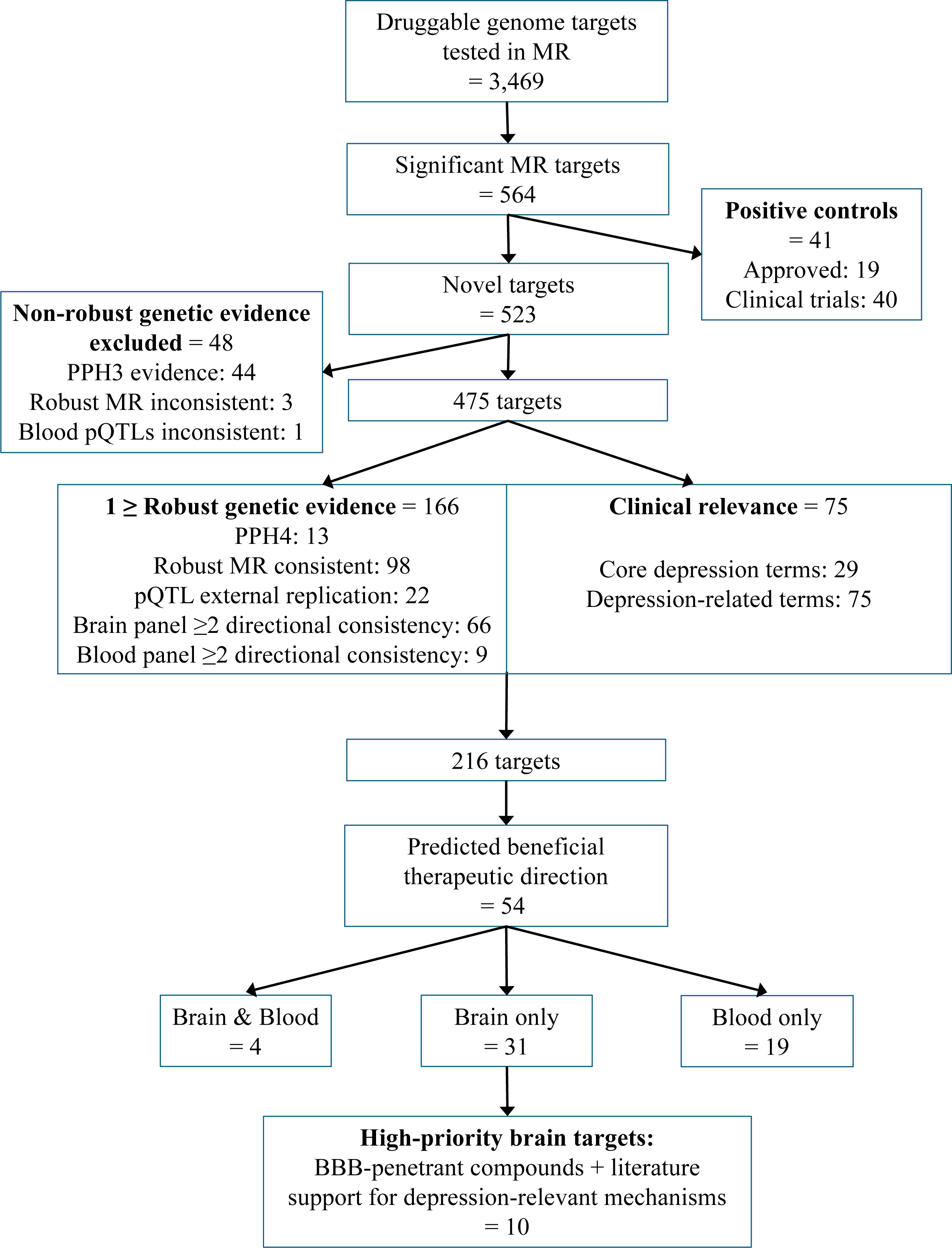
Novel target identification and prioritisation workflow. Flow diagram showing systematic identification of 10 high-priority novel drug targets for major depressive disorder from 3,469 druggable genome targets tested in cis-Mendelian randomisation.

#### Genetic robustness assessment

To evaluate genetic evidence, we incorporated four assessments. First, we favoured targets with consistent directional *cis*-MR effects across ≥2 discovery panels within the same tissue type (brain or blood). We excluded blood pQTL targets with directionally opposite *cis*-MR effects between discovery panels using different protein assay platforms, indicating technical artefacts in protein measurement (Supplementary Methods). Second, we favoured targets with robust MR estimates that aligned with the primary MR-IVW effect direction and excluded those with directional disagreement. Third, we prioritised targets with PPH4 colocalisation evidence, excluding those with PPH3 evidence. Fourth, we prioritised pQTL targets with external replication. Not all targets had sufficient data or statistical power to be evaluated by each of these criteria. We therefore prioritised targets meeting ≥1 of these criteria, alongside therapeutic potential.

#### Therapeutic potential assessment

We focused on identifying repurposing candidates, defined as targets of drugs approved for the treatment of non-psychiatric conditions. We also considered targets of compounds tested in clinical trials for indications other than depression or anxiety. We evaluated clinical relevance by identifying targets with compounds developed for other conditions that showed depression-related drug effects. We integrated our therapeutic beneficial compound analysis by prioritising targets with at least one compound that has mechanisms acting in a direction that would oppose the target risk-increasing effect indicated by our MR results.

#### Final prioritisation criteria

Among significant targets from our primary *cis*-MR analysis, we prioritised targets meeting two criteria: 1) either ≥1 genetic robustness evidence or targeted by compounds showing depression-related drug effects, and 2) targeted by ≥1 compound(s) with estimated beneficial therapeutic direction. We then categorised prioritised targets by tissue expression (brain-only, blood-only or brain and blood). We investigated brain-only targets as high-priority given the neurobiological basis of MDD and that tissue-specific targets may produce fewer off-target effects than those ubiquitously expressed [41]. Among prioritised brain-only targets, we further defined high-priority targets as those meeting both of the following criteria through literature searches: 1) blood-brain barrier permeability of estimated beneficial compounds, and 2) the target’s role in depression-relevant pathophysiological mechanisms. This included literature on the target function in relevant biological processes and evidence for antidepressant-like effects of beneficial compounds or target modulation in animal models or human studies.

#### Positive control analysis

We identified positive controls (PCs), also known as genetic rediscoveries of known drug-target mechanisms, using drug effects data from ChEMBL and databases of drugs licensed in the UK [28]. We selected targets of drugs with primary indications for depression and/or anxiety and classified them as approved PC targets (our primary positive controls, targeted by drugs in phase IV) and clinical trial PC targets (our secondary positive controls, targeted by compounds in clinical trial phases I-III). We included anxiety due to the high comorbidity, treatment and genetic overlap between anxiety disorders and MDD, with genetic correlations approaching unity (∼0.9) [42–44]. As a single compound can act on multiple targets, we also analysed our results at the compound level (Supplementary Materials). We conducted enrichment analyses to assess whether our *cis*-MR approach identified known drug targets for depression. We separately compared significance rates in brain and blood tissues of 1) approved PC targets versus all other targets, and 2) clinical trial PC targets versus all other non-PC targets. An odds ratio (OR) > 1 indicated higher odds of PC targets among significant results (Supplementary Methods).

## Results

### Overview

Our primary *cis*-MR analysis identified 564 unique drug targets (of 3,469 tested) significantly associated with MDD across at least one of ten panels of protein or transcript values measured in blood, brain, and CSF (see Supplementary Tables 4-13 for genetic results in each panel). We evaluated significant targets by integrating evidence for genetic robustness and therapeutic potential (**Figure 1** and Supplementary Table 14). Target-level aggregated results from all analyses for each of the 564 significant targets are in Supplementary Table 15.

### Positive control (PC) analysis

Our *cis*-MR analysis demonstrated stronger genetic support for compounds in clinical trial phase IV for depression and anxiety (approved positive controls (PC)) than those in clinical trial I-III (clinical trial PCs). At the drug level, 50/61 (82%) approved PCs showed at least one significant MR target effect on MDD, compared to 86/169 (51%) for clinical trial PC compounds (Supplementary Tables 16-18). Of the approved PC drugs with significant targets, most showed estimated beneficial mechanisms of action. Specifically, 27 drugs (54%) had targets with exclusively beneficial effects (where drug mechanisms oppose target MDD risk directions indicated by MR), while 13 drugs (26%) with multiple targets showed mixed effects (i.e. beneficial on some targets, adverse on others). Only 4 (8%) exhibited exclusively adverse effects, and 6 (12%) had unknown directionality. For compounds undergoing clinical trials, 29 (33.7%) showed beneficial mechanisms, 24 (27.9%) had mixed effects, 15 (17.4%) demonstrated adverse effects, and 18 (20.9%) had unknown directionality. **Table 1** presents results for targets of key positive controls with significant MR effects and estimated therapeutic directions that align with their known clinical efficacy for MDD and/or anxiety.

**Table 1:** Drug targets of positive control compounds for depression and anxiety with significant MR effects and estimated beneficial therapeutic directions on MDD.

| Target | Tissue (MR effect direction) | Supporting genetic evidence | Estimated therapeutic MOA & drug MOA | Compounds with aligned MOA | Primary indications |
| --- | --- | --- | --- | --- | --- |
| **Monoamine signalling** |  |  |  |  |  |
| Sodium-dependent serotonin transporter:<br>*SLC6A4* | eQTL blood (+) | Effect direction aligned with beneficial MOA therapeutic effects | Inhibition (-),<br>28 inhibitors/<br>antagonists (-) | 21 SSRIs/SNRIs,<br>TCAs, atypical<br>antidepressants | Depression & anxiety |
| Sodium-dependent dopamine transporter:<br>*SLC6A3* | eQTL cortex (+) | Effect direction aligned with beneficial MOA therapeutic effects | Inhibition (-),<br>10 inhibitors (-) | bupropion | Depression,<br>ADHD (e.g.<br>methylphenidate) |
| Dopamine receptor D2:<br>*DRD2* | eQTLs (+): basal ganglia,<br>hippocampus, spinal cord | Directional consistency, aligned with beneficial MOA therapeutic effects | Inhibition (-),<br>46 antagonists/<br>inhibitors (-) | amoxapine,<br>quetiapine<br>prochlorperazine | Depression, anxiety,<br>bipolar disorder,<br>schizophrenia |
| Histamine H1 receptor:<br>*HRH1* | eQTL cortex (+),<br>eQTL blood (+) | Directional consistency, aligned with beneficial MOA therapeutic effects | Inhibition (-),<br>62 antagonists/<br>inverse agonists (-) | hydroxyzine,<br>dibenzepin | Depression & anxiety |
| **Glutamatergic signalling** |  |  |  |  |  |
| NMDA receptor subunits:<br>GRIN1 (+)<br>GRIN2D (+)<br>GRIN2A (-) | pQTL DLPFC (+),<br>eQTL cerebellum (-),<br>pQTL blood (+) | Largely consistent effect across subunits (+) aligned with beneficial MOA therapeutic effects | Inhibition/<br>activation (-/+),<br>21 negative allosteric modulator/antagonists/<br>blocker (-) | esketamine | Treatment-resistant depression |

| Target | Tissue (MR effect direction) | Supporting genetic evidence | Estimated therapeutic MOA & drug MOA | Compounds with aligned MOA | Primary indications |
| --- | --- | --- | --- | --- | --- |
| **GABA Signalling** |  |  |  |  |  |
| GABA(A) receptor subunits: *GABRA6* (-)<br>*GABRB2* (-)<br>*GABRG2* (-),<br>*GABRP* (+) | eQTL cerebellum (-/+) | Largely Consistent effect across subunits (-) aligned with beneficial MOA therapeutic effects of anxiolytics | Inhibition/activation (-/+),<br>59 positive allosteric modulators/agonist (+) | benzodiazepines, brexanolone, meprobamate | Anxiety, insomnia, anxiety with depressive symptoms, postpartum depression |
| **Calcium Ion Channel Modulators** |  |  |  |  |  |
| Calcium channel subunits: *CACNG2* (+),<br>*CACNA1G* (+),<br>*CACNA2D2* (+/-)<br>*CACNA2D4* (-) | eQTLs (+/-): cerebellum, cortex, basal ganglia, hippocampus, blood | Largely consistent effect across subunits (+), robust MR support | Inhibition/activation (-/+), modulators, blockers (-) | pregabalin | Generalised anxiety disorder, epilepsy, nerve pain |
| **Targets of compounds in clinical trials for depression/anxiety** |  |  |  |  |  |
| Phosphodiesterases: *PDE6B* (+),<br>*PDE6G* (+),<br>*PDE6H* (+)<br>*PDE3B* (+)<br>*PDE6C* (-) | eQTL blood (+/-) | Largely consistent effect across subtypes (+) aligned with beneficial MOA | Inhibition/activation (-/+), inhibitors (-) | 9 inhibitors, e.g. pentoxifylline dipyridamole | Cardiovascular (IV), depression (I-II) |
| Corticotropin-releasing factor receptor 1: *CRHR1* | eQTLs: cerebellum (-), cortex (+) | Cortex effect (+) aligned with beneficial MOA, robust MR support | Inhibition/activation (-/+), 5 antagonists (-) | 5 in clinical trial: e.g. verucerfont, emicerfont, pexacerfont | Depression & anxiety (I-II) |

*Note: A positive (+) MR effect indicates higher expression increases MDD risk, suggesting beneficial compounds would have inhibitory (-) mechanisms of action (MOA) on the target; a negative (-) MR effect indicates lower expression increases risk, suggesting beneficial compounds would have activating (+) mechanisms of action. Consistent MR effect directions across related subunits/subtypes, concordance between tissues, and agreement between genetically estimated therapeutic direction and known clinical efficacy of established drug mechanisms strengthen the validity of our findings. ADHD, attention-deficit/hyperactivity disorder; DLPFC, dorsolateral prefrontal cortex; eQTL, expression quantitative trait loci; MDD, major depressive disorder; MOA, mechanism of action; MR, Mendelian randomisation; pQTL, protein quantitative trait loci.*

At the target-level, we found significant enrichment of targets of approved PC compounds in brain tissues (OR=1.97, 95% CI=1.11-3.49, p=0.018; Supplementary Table 19). Approved PC targets had a 1.7-fold higher rate of significant MR estimates (18.8%) in the brain compared to other targets (10.5%). Notably, the aforementioned compound-level rate (82%) was higher because most drugs act on multiple targets, and compounds required only one of those targets to be significant to be counted. Targets of clinical trial PCs showed similar brain-specific enrichment (OR=1.54, 95% CI=1.02-2.30, p=0.036). Enrichment was not significant in blood tissues for either approved PC targets (OR=1.81, 95% CI=0.75-4.35, p=0.289) or clinical trial PC targets (OR=1.10, 95% CI=0.60-2.03, p=0.756). *Cis*-MR evidence for positive controls in brain tissues supports the biological validity of our approach, aligning with the neurobiological aetiology of MDD and with established pharmacological interventions targeting neurotransmitter systems and neural circuitry [45].

### Prioritisation of novel targets for repurposing

We identified 523 drug targets beyond those currently used or explored in depression treatment. Following our systematic prioritisation framework (**Figure 1**), 216 targets met criteria for genetic robustness evidence or clinical relevance, of which 54 were prioritised due to being targeted by compounds with estimated beneficial therapeutic directions for MDD (Supplementary Table 15). Tissue expression patterns of prioritised targets included four found in both brain and blood panels, 31 brain-only targets, and 19 blood-only targets. Of the 31 targets exhibiting brain-specific effects, 10 met our high-priority criteria with blood-brain barrier-penetrant compounds and supporting literature for depression-relevant mechanisms (**Table 2** and Supplementary Table 20).

**Table 2:** High-priority brain targets of novel compounds with estimated beneficial therapeutic directions on MDD.

| Target | Tissue (MR effect) | Supporting genetic evidence | Estimated therapeutic MOA & available drug MOA | Beneficial compounds | Primary indications (depression-related side effects/ clinical trial phase) | Depression relevant mechanisms |
| --- | --- | --- | --- | --- | --- | --- |
| Cyclin-dependent kinase 4: *CDK4* | eQTLs: basal ganglia (-), cerebellum (-), hippocampus (-), spinal cord (-), pQTL DLPFC (+) | Directional consistency eQTL brain, external replication pQTL | Mixed: inhibition/ activation (-/+), inhibitor (-) | Crosses BBB: abemaciclib | Breast cancer (fatigue, appetite, psychiatric) | Neuroplasticity, neural activity, neurogenesis |
| Nischarin Imidazoline receptor 1: *NISCH* | eQTLs: cerebellum (+), hippocampus (+), spinal cord (-) | Directional consistency cerebellum & hippocampus | Mixed: inhibition/ activation (-/+), agonist (+) | Crosses BBB: moxonidine, rilmenidine | Hypertension (insomnia, anxiety, depression, psychiatric) | Sympathetic regulation, stress response, target activation has animal model antidepressant-like effects |
| Nitric oxide synthase, inducible: *NOS2* | eQTLs: cortex (+), basal ganglia (+), cerebellum (+) | Directional consistency | Inhibition (-), inhibitor (-) | Clinical trial: tilarginine, KD7040, GW-274150; crosses BBB: BN-4 | Clinical trial: cardiogenic shock (III), migraine (II) | Neuroinflammation, oxidative stress, target inhibition has animal model antidepressant-like effects |
| Glutaminase kidney isoform, mitochondrial: *GLS* | eQTLs: cortex (+), cerebellum (+) | Directional consistency | Inhibition (-), inhibitor (-) | Clinical trial: telaglenastat; crosses BBB: JHU-083 | Clinical trial: cancer (II) | Neuroinflammation, glutamate signalling, target inhibition has animal model antidepressant-like effects |
| Neurotrophic receptor tyrosine kinase 3: *NTRK3* | eQTL cortex (+) | Colocalisation | Inhibition (-), inhibitor (-) | Crosses BBB: entrectinib, larotrectinib | Cancer (anxiety, depression, psychiatric, mood, appetite/weight, fatigue/sleep) | Neuroplasticity, monoamine signalling, HPA axis regulation |
| Angiotensin-converting enzyme: *ACE* | eQTL cortex (+) | MR effect direction aligns with drug MOA & reported mental health benefits | Inhibition (-), inhibitor (-) | Crosses BBB: e.g. captopril, fosinopril, lisinopril | Hypertension (anxiety, depression, mood, appetite/weight, insomnia/sleep) | Neuroinflammation, synaptic plasticity, serotonergic signalling, stress response, ACE inhibitors have antidepressant-like effects in animal models and clinical studies |

| Target | Tissue (MR effect) | Supporting genetic evidence | Estimated therapeutic MOA & available drug MOA | Beneficial compounds | Primary indications (depression-related side effects/ clinical trial phase) | Depression relevant mechanisms |
| --- | --- | --- | --- | --- | --- | --- |
| Complex I NADH dehydrogenase subunits: *NDUFA2* (+) *NDUFB6* (+) *NDUFS1* (+) | eQTL cortex (+) | Consistent effect across subunits, robust MR support, reported mental health benefits | Inhibition (-), inhibitor (-) | Crosses BBB: metformin | Type 2 diabetes (appetite) | Mitochondrial function, neuroinflammation, metformin has antidepressant-like effects in animal models and human observational studies |
| Hepatocyte growth factor receptor: *MET* | eQTLs: basal ganglia (+), spinal cord (+) | Directional consistency | Inhibition (-), inhibitor (-) | Crosses BBB: capmatinib, cabozantinib | Cancer (appetite, fatigue, psychiatric) | Neuroplasticity, neurotrophic signalling, neuroinflammation |

*Note: Prioritised targets with novel repurposing opportunities for MDD, i.e. drugs already approved or > Phase I for other indications but not in clinical trial for depression/anxiety as a primary indication. A positive (+) MR effect indicates higher expression increases MDD risk, suggesting beneficial compounds would have inhibitory (-) mechanisms of action (MOA) on the target; a negative (-) MR effect indicates lower expression increases risk, suggesting beneficial compounds would have activating (+) mechanisms of action. Brain targets were prioritised based on: availability of compounds acting in the beneficial direction for at least one tissue, plus clinical relevance (depression-related side effects) or ≥ 1 robust genetic evidence (significant MR effects with directional consistency in multiple brain/nervous system tissues, robust MR support, colocalisation support or pQTL external replication), plus compounds with blood-brain-barrier permeability and plausible depression-relevant biological mechanisms including pre-clinical evidence. BBB, blood-brain barrier; DLPFC, dorsolateral prefrontal cortex; eQTL, expression quantitative trait loci; MDD, major depressive disorder; MOA, mechanism of action; MR, Mendelian randomisation; pQTL, protein quantitative trait loci.*

## Discussion

We identified novel drug targets and immediate repurposing opportunities for MDD by triangulating genetic evidence and clinical data. Our integrated framework strengthens causal inference in target identification, guiding repurposing. The target-compound pairs we prioritise span MDD drug development stages. These include long-standing antidepressants, recently approved treatments, repurposing compounds in MDD clinical trials or with early clinical evidence not yet formally tested for MDD treatment, and candidates with strong preclinical evidence.

### Genetic validation through established targets and mechanisms

Our findings suggest genetic evidence informs clinical success in MDD drug development, with a higher rate of significant targets for phase IV positive control drugs (82%) versus earlier phases I-III compounds (51%). We corroborate broader findings that genetically supported compounds for complex traits are more likely to be approved [7]. Additionally, targets of positive control compounds showed enrichment in brain tissues, validating our focus on brain targets for repurposing.

For monoaminergic targets of approved MDD treatments, risk-increasing expression of *SLC6A4* (encoding the serotonin transporter), *SLC6A3* (dopamine transporter), and *DRD2* (dopamine D2 receptor) aligned with the established therapeutic inhibitory mechanisms of antidepressants, bupropion, and antipsychotics, respectively. Genetic evidence for DRD2 antagonism was consistent across brain regions, including the basal ganglia, aligning with its role in reward processing [46]. By specifically implicating DRD2 antagonism, we provide directional insight into the PGC MDD-3 GWAS findings that identified the *DRD2* gene as the strongest gene-based association and antipsychotic compound enrichment [16]. Beyond monoaminergic signalling in MDD, we also found genetic support for anxiety medications, reflecting the high comorbidity and genetic overlap between these disorders [42–44]. For example, multiple calcium channel subunits showed significant effects in the brain, aligning with pregabalin’s efficacy in depression-anxiety comorbidity [47], and the PGC MDD-3 finding that pregabalin was among the top-enriched compounds [16].

Positive control compounds with multiple targets showed both exclusively beneficial and mixed therapeutic effects. For the dual-action tricyclic antidepressant Amoxapine, genetic effects aligned with therapeutic benefits for both serotonergic (via SLC6A4 inhibition) and dopaminergic (via DRD2 antagonism) signalling. Conversely, for esketamine, genetic effects aligned with therapeutic benefits of inhibiting GRIN1 and GRIN2D, but adverse for GRIN2A, consistent with mixed therapeutic and side effect profiles for different NMDA receptor subunits [48,49]. Similar patterns were observed among GABAA receptor subunits targeted by anxiolytic benzodiazepines. Target-specific genetic effects suggest which receptor subtypes may drive therapeutic benefits versus contribute to side effects [48–50]. Our findings could potentially inform the development of more selective compounds with improved efficacy-to-side-effect profiles for MDD.

### Cardiovascular and metabolic repurposing compounds

Our genetic evidence supports repurposing of multiple types of cardiovascular and metabolic system drugs for MDD, from different stages in the repurposing pipeline. For pentoxifylline, a PDE inhibitor approved for vascular disease and tested in depression clinical trials [51–54], we found four beneficial PDE6 and PDE3 subunits and one adverse, guiding subtype-specific targeting to improve efficacy [55]. A meta-analysis of Phase II trials found adjunctive pentoxifylline significantly improved depressive symptoms [56]. Among compounds with preliminary clinical evidence and yet to be formally evaluated in MDD clinical trials, *ACE* gene expression showed a risk-increasing effect in cortical tissue. BBB-penetrant ACE inhibitors like captopril used in the treatment of hypertension have demonstrated animal model antidepressant-like effects and mental health benefits as a secondary outcome in clinical trials [57,58] potentially by reducing oxidative stress and neuroinflammation while enhancing serotonergic signalling [59]. Both PDE inhibitors and ACE inhibitors are widely used and well-tolerated in cardiovascular medicine, adding to their potential for repurposing.

We identified risk-increasing effects on MDD for Complex I subunit cortical expression (*NDUFA2*, *NDUFB6*, *NDUFS1* genes), aligning with the inhibitory mechanism of metformin on these targets, a type 2 diabetes medication. This complements the identification of another metformin gene encoding target, *NDUFAF3*, as a novel MDD-associated gene [60]. Metformin exhibits antidepressant and anxiolytic-like effects in animal models [61,62] and is associated with a decreased risk of depression in observational case-control studies [63]. Whilst one phase III trial demonstrated metformin’s efficacy in treatment-resistant bipolar depression [64], the only published clinical trial formally investigating metformin for MDD treatment as a primary outcome was retracted [65]. Given our genetic evidence alongside preclinical and observational support, prioritising metformin for clinical trials with MDD as a primary outcome is warranted.

Preclinical evidence supports the potential of antihypertensive imidazoline I_1_ agonists for MDD. We found mixed effects of *NISCH* (encoding Nischarin, the proposed I_1_ receptor) across brain regions, likely reflecting context-dependent roles in neuroinflammation and neural plasticity [66,67]. I_1_ agonist agmatine may be an endogenous antidepressant [68], with compensatory increased brain levels during stress and antidepressant-like effects in rodents [69–71] alongside *NISCH* amygdala downregulation [72]. SSRI efficacy may be partially mediated by I_1_ activation [73,74]. Consistent with our risk-increasing brain effects, Nischarin was elevated in postmortem prefrontal cortex of MDD patients but lower in those taking antidepressants [75]. These findings establish Nischarin as a relevant MDD target through potential involvement in stress-depression pathways and antidepressant action.

### Cancer repurposing compounds

Among our prioritised novel targets were three proteins inhibited by BBB-penetrant cancer drugs, with plausible roles in neural function. CDK4 showed mixed directional effects across five brain regions, reflecting diverse neural functions regulating neurogenesis [76], neuronal activity [77] and synaptic plasticity [78]. *NTRK3* encodes the receptor for neurotrophin-3, which modulates monoaminergic transmission, BDNF expression and HPA axis function [79]. We corroborate the PGC MDD-3 identification of *NTRK3* as a high-confidence MDD gene [16] whilst providing novel directional insights by identifying inhibitors to counteract the risk-increasing effect of NTRK3. *MET* encodes the MET hepatocyte growth factor receptor, involved in regulating neuroplasticity, neurotrophic pathways, and inflammation [80]. Approved inhibitors for all three targets exhibit psychiatric side effects, supporting their neuropsychiatric relevance [81–83]. However, the CDK4-selective inhibitor abemaciclib demonstrates the potential for an improved nervous system safety profile when repurposing cancer drugs for MDD, with fewer psychiatric side effects than less selective alternatives [83]. Further investigations of CDK4, NTRK3 and MET inhibitors in MDD are warranted, such as dose-ranging studies or selective compound development to optimise efficacy while minimising side effects.

### Early-stage drug development compounds

We identified two enzyme targets inhibited by experimental compounds yet to be clinically approved for any indication or tested in MDD clinical trials, representing opportunities beyond the repurposing of approved medications. Inhibiting GLS (glutaminase) offers a mechanistically distinct approach to glutamatergic modulation compared to approved depression compounds. While NMDA antagonists (e.g. esketamine) block glutamate receptor signalling, glutaminase inhibition reduces glutamate production in activated microglia, directly limiting excitotoxicity and neuroinflammation [84,85]. The novel BBB-penetrant glutaminase inhibitor JHU-083 demonstrates antidepressant-like effects in animal models while minimising peripheral side effects as a pro-drug [84,85]. Similarly, inhibition of inducible nitric oxide synthase (iNOS, encoded by *NOS2*) targets glial-induced neuroinflammation [86,87]. Unlike other NOS isoforms, iNOS offers temporal and cell-type-specificity to reduce neuroinflammation without affecting normal NO signalling [86,87]. Selective iNOS inhibition in rodents exhibits antidepressant-like effects [88,89] and attenuates stress-induced anxiogenic effects [90]. To our knowledge, we provide the first human genomic evidence for these preclinical compounds, highlighting the potential for novel MDD therapeutics targeting neuroinflammatory pathways.

### Limitations and future directions

Whilst we prioritised drugged targets for rapid translation and included high-quality experimental targets beyond prior studies [17,18], other pQTL/eQTLs may in the future become therapeutically viable [9]. Genetic effect directions not aligning with known therapeutic mechanisms may reflect differences between lifelong genetic versus acute pharmacological effects [91], tissue [10] or cell-specific effects [29]. Furthermore, c*is*-MR examines the effects of individual proteins, however, proteins function within complex pathways [92], challenging estimation of therapeutic effects from single-target modulation. Polypharmacological effects can trigger broader network responses and compensatory mechanisms [92,93]. While *cis*-MR reduces the risk of horizontal pleiotropy and we applied robust MR estimators, it cannot be entirely ruled out [10]. Only 11% of targets had sufficient power for colocalisation, which requires strong *cis*-region GWAS associations in both traits [94]. Our analysis of European-associated ancestry data may limit generalisability to other populations [95]. Experimental validation and clinical studies are needed to confirm the therapeutic potential of novel targets, with research required in globally representative populations and diverse biological contexts.

MDD heterogeneity challenges translating genetically supported targets and compounds into treatments. Despite genetic evidence from our study and previous work [18], CRHR1 antagonists targeting stress-dysregulation have failed in depression clinical trials [96]. Similarly, anti-inflammatory drug clinical trials have largely been unsuccessful [97]. These failures may stem from a ‘one-size fits all’ approach, obscuring therapeutic effects for a subset of patients [97,98]. Future work could explore genetically supported compounds through patient stratification, matching treatments to MDD subtypes e.g., stress-dysregulation or inflammation-driven symptoms [97,98].

### Conclusions

Our genetically informed approach corroborated established MDD targets while identifying novel therapeutic opportunities beyond traditional monoaminergic mechanisms. Several novel targets have approved drugs for other indications with established safety profiles and BBB-penetration, offering opportunities for immediate clinical evaluation.

## Acknowledgements

This project has received funding from the European Research Council (ERC) under the European Union’s Horizon 2020 research and innovation programme (I-IRISK grant agreement No. 863981). AFS is supported by BHF grants PG/18/5033837, PG/22/10989, the UCL BHF Research Accelerator AA/18/6/34223. AFS received additional support from the National Institute for Health Research University College London Hospitals Biomedical Research Centre. SB is supported by the Great Ormond Street Hospital Children’s Charity. This work was funded by UK Research and Innovation (UKRI) under the UK government’s Horizon Europe funding guarantee EP/Z000211/1, by the UKRI/NIHR Multimorbidity fund Mechanism and Therapeutics Research Collaborative MR/V033867/1, and by the Rosetrees Trust. The authors acknowledge the use of the UCL Myriad High Performance Computing Facility (Myriad@UCL), and associated support services, in the completion of this work. This research has been conducted using the UK Biobank Resource under application number 12113. We are grateful to the UK Biobank participants. The UK Biobank was established by the Wellcome Trust medical charity, Medical Research Council, Department of Health, Scottish Government, and the Northwest Regional Development Agency. It has also had funding from the Welsh Assembly Government and the British Heart Foundation. We thank the research participants and employees of 23andMe, Inc. for their contribution to the GWAS summary data used in this study.

## Ethical approval

The UK Biobank has ethical approval from the North West Multi-centre Research Ethics Committee as a Research Tissue Bank approval (REC reference: 21/NW/0157).

## Data availability

GWAS summary statistics for MDD are available from the Psychiatric Genomics Consortium at https://pgc.unc.edu/for-researchers/download-results/. GWAS summary statistics including 23andMe data require an approved application through 23andMe available to qualified researchers under an agreement with 23andMe that protects the privacy of the 23andMe participants (visit https://research.23andme.com/dataset-access/). UK Biobank data are available through application at https://www.ukbiobank.ac.uk/enable-your-research/apply-for-access/. Open Targets tractability data can be downloaded from http://ftp.ebi.ac.uk/pub/databases/opentargets/platform/latest/input/target/tractability/. QTL datasets are available from: Blood plasma pQTL data: deCODE (https://www.decode.com/summarydata/), UKB-PPP (https://www.synapse.org/#!Synapse:syn51364943/), INTERVAL (http://www.phpc.cam.ac.uk/ceu/proteins/), and Gudjonsson (https://www.ebi.ac.uk/gwas/publications/35078996). Brain pQTL data: ROSMAP and Banner (https://www.synapse.org/#!Synapse:syn24172458). CSF pQTL data: Yang (https://dss.niagads.org/datasets/ng00102/). Blood eQTL data: eQTLGen (https://www.eqtlgen.org/). Brain eQTL data: MetaBrain (https://www.metabrain.nl/).

## References

1. GBD 2019 Mental Disorders Collaborators. Global, regional, and national burden of 12 mental disorders in 204 countries and territories, 1990-2019: a systematic analysis for the Global Burden of Disease Study 2019. Lancet Psychiatry. 2022;9:137–50.

2. Trivedi MH, Rush AJ, Wisniewski SR, Nierenberg AA, Warden D, Ritz L, et al. Evaluation of outcomes with citalopram for depression using measurement-based care in STAR*D: implications for clinical practice. Am J Psychiatry. 2006;163:28–40.

3. Souery D, Serretti A, Calati R, Oswald P, Massat I, Konstantinidis A, et al. Switching antidepressant class does not improve response or remission in treatment-resistant depression. J Clin Psychopharmacol. 2011;31:512–6.

4. Harmer CJ, Duman RS, Cowen PJ. How do antidepressants work? New perspectives for refining future treatment approaches. Lancet Psychiatry. 2017;4:409–18.

5. King EA, Davis JW, Degner JF. Are drug targets with genetic support twice as likely to be approved? Revised estimates of the impact of genetic support for drug mechanisms on the probability of drug approval. PLoS Genet. 2019;15:e1008489.

6. Nelson MR, Tipney H, Painter JL, Shen J, Nicoletti P, Shen Y, et al. The support of human genetic evidence for approved drug indications. Nat Genet. 2015;47:856–60.

7. Minikel EV, Painter JL, Dong CC, Nelson MR. Refining the impact of genetic evidence on clinical success. Nature. 2024;629:624–9.

8. Rusina PV, Falaguera MJ, Romero JMR, McDonagh EM, Dunham I, Ochoa D. Genetic support for FDA-approved drugs over the past decade. Nat Rev Drug Discov. 2023;22:864.

9. Finan C, Gaulton A, Kruger FA, Lumbers RT, Shah T, Engmann J, et al. The druggable genome and support for target identification and validation in drug development. Sci Transl Med [Internet]. 2017;9. Available from: 10.1126/scitranslmed.aag1166

10. Schmidt AF, Finan C, Gordillo-Marañón M, Asselbergs FW, Freitag DF, Patel RS, et al. Genetic drug target validation using Mendelian randomisation. Nat Commun. 2020;11:3255.

11. Hingorani AD, Kuan V, Finan C, Kruger FA, Gaulton A, Chopade S, et al. Improving the odds of drug development success through human genomics: modelling study. Sci Rep. 2019;9:18911.

12. Daghlas I, Gill D. Mendelian randomization as a tool to inform drug development using human genetics. Cambridge Prisms: Precision Medicine. 2023;1:e16.

13. Walker VM, Davey Smith G, Davies NM, Martin RM. Mendelian randomization: a novel approach for the prediction of adverse drug events and drug repurposing opportunities. Int J Epidemiol. 2017;46:2078–89.

14. Gordillo-Marañón M, Zwierzyna M, Charoen P, Drenos F, Chopade S, Shah T, et al. Validation of lipid-related therapeutic targets for coronary heart disease prevention using human genetics. Nat Commun. 2021;12:6120.

15. Storm CS, Kia DA, Almramhi MM, Bandres-Ciga S, Finan C, International Parkinson’s Disease Genomics Consortium (IPDGC), et al. Finding genetically-supported drug targets for Parkinson’s disease using Mendelian randomization of the druggable genome. Nat Commun. 2021;12:7342.

16. Major Depressive Disorder Working Group of the Psychiatric Genomics Consortium. Trans-ancestry genome-wide study of depression identifies 697 associations implicating cell types and pharmacotherapies. Cell [Internet]. 2025; Available from: 10.1016/j.cell.2024.12.002

17. Liu J, Cheng Y, Li M, Zhang Z, Li T, Luo X-J. Genome-wide Mendelian randomization identifies actionable novel drug targets for psychiatric disorders. Neuropsychopharmacology. 2023;48:270–80.

18. Li Y, Dang X, Chen R, Teng Z, Wang J, Li S, et al. Cross-ancestry genome-wide association study and systems-level integrative analyses implicate new risk genes and therapeutic targets for depression. Nat Hum Behav. 2025;9:806–23.

19. Schmidt AF, Bourfiss M, Alasiri A, Puyol-Anton E, Chopade S, van Vugt M, et al. Druggable proteins influencing cardiac structure and function: Implications for heart failure therapies and cancer cardiotoxicity. Sci Adv. 2023;9:eadd4984.

20. Ferkingstad E, Sulem P, Atlason BA, Sveinbjornsson G, Magnusson MI, Styrmisdottir EL, et al. Large-scale integration of the plasma proteome with genetics and disease. Nat Genet. 2021;53:1712–21.

21. Sun BB, Chiou J, Traylor M, Benner C, Hsu Y-H, Richardson TG, et al. Plasma proteomic associations with genetics and health in the UK Biobank. Nature. 2023;622:329– 38.

22. Võsa U, Claringbould A, Westra H-J, Bonder MJ, Deelen P, Zeng B, et al. Large-scale cis- and trans-eQTL analyses identify thousands of genetic loci and polygenic scores that regulate blood gene expression. Nat Genet. 2021;53:1300–10.

23. Yang C, Farias FHG, Ibanez L, Suhy A, Sadler B, Fernandez MV, et al. Genomic atlas of the proteome from brain, CSF and plasma prioritizes proteins implicated in neurological disorders. Nat Neurosci. 2021;24:1302–12.

24. Robins C, Liu Y, Fan W, Duong DM, Meigs J, Harerimana NV, et al. Genetic control of the human brain proteome. Am J Hum Genet. 2021;108:400–10.

25. de Klein N, Tsai EA, Vochteloo M, Baird D, Huang Y, Chen C-Y, et al. Brain expression quantitative trait locus and network analyses reveal downstream effects and putative drivers for brain-related diseases. Nat Genet. 2023;55:377–88.

26. Eldjarn GH, Ferkingstad E, Lund SH, Helgason H, Magnusson OT, Gunnarsdottir K, et al. Large-scale plasma proteomics comparisons through genetics and disease associations. Nature. 2023;622:348–58.

27. Brown KK, Hann MM, Lakdawala AS, Santos R, Thomas PJ, Todd K. Approaches to target tractability assessment - a practical perspective. Medchemcomm. 2018;9:606–13.

28. Mendez D, Gaulton A, Bento AP, Chambers J, De Veij M, Félix E, et al. ChEMBL: towards direct deposition of bioassay data. Nucleic Acids Res. 2019;47:D930–40.

29. Hukerikar N, Hingorani AD, Asselbergs FW, Finan C, Schmidt AF. Prioritising genetic findings for drug target identification and validation. Atherosclerosis. 2024;390:117462.

30. Bycroft C, Freeman C, Petkova D, Band G, Elliott LT, Sharp K, et al. The UK Biobank resource with deep phenotyping and genomic data. Nature. 2018;562:203–9.

31. Burgess S, Zuber V, Valdes-Marquez E, Sun BB, Hopewell JC. Mendelian randomization with fine-mapped genetic data: Choosing from large numbers of correlated instrumental variables. Genet Epidemiol. 2017;41:714–25.

32. Bowden J, Del Greco M F, Minelli C, Davey Smith G, Sheehan N, Thompson J. A framework for the investigation of pleiotropy in two-sample summary data Mendelian randomization. Stat Med. 2017;36:1783–802.

33. Burgess S, Davey Smith G, Davies NM, Dudbridge F, Gill D, Glymour MM, et al. Guidelines for performing Mendelian randomization investigations. Wellcome Open Res. 2019;4:186.

34. Patel A, Ye T, Xue H, Lin Z, Xu S, Woolf B, et al. MendelianRandomization v0.9.0: updates to an R package for performing Mendelian randomization analyses using summarized data. Wellcome Open Res. 2023;8:449.

35. Slob EAW, Burgess S. A comparison of robust Mendelian randomization methods using summary data. Genet Epidemiol. 2020;44:313–29.

36. Burgess S, Thompson SG. Mendelian Randomization: Methods for Using Genetic Variants in Causal Estimation. CRC Press; 2015.

37. Burgess S, Foley CN, Allara E, Staley JR, Howson JMM. A robust and efficient method for Mendelian randomization with hundreds of genetic variants. Nat Commun. 2020;11:376.

38. Bowden J, Davey Smith G, Haycock PC, Burgess S. Consistent Estimation in Mendelian Randomization with Some Invalid Instruments Using a Weighted Median Estimator. Genet Epidemiol. 2016;40:304–14.

39. Wallace C. Eliciting priors and relaxing the single causal variant assumption in colocalisation analyses. PLoS Genet. 2020;16:e1008720.

40. Zuber V, Grinberg NF, Gill D, Manipur I, Slob EAW, Patel A, et al. Combining evidence from Mendelian randomization and colocalization: Review and comparison of approaches. Am J Hum Genet. 2022;109:767–82.

41. Duffy Á, Verbanck M, Dobbyn A, Won H-H, Rein JL, Forrest IS, et al. Tissue-specific genetic features inform prediction of drug side effects in clinical trials. Sci Adv. 2020;6:eabb6242.

42. Strom NI, Verhulst B, Bacanu S-A, Cheesman R, Purves KL, Gedik H, et al. Genome-wide association study of major anxiety disorders in 122,341 European-ancestry cases identifies 58 loci and highlights GABAergic signaling [Internet]. medRxiv. 2024. Available from: http://medrxiv.org/lookup/doi/10.1101/2024.07.03.24309466

43. Kessler RC, Sampson NA, Berglund P, Gruber MJ, Al-Hamzawi A, Andrade L, et al. Anxious and non-anxious major depressive disorder in the World Health Organization World Mental Health Surveys. Epidemiol Psychiatr Sci. 2015;24:210–26.

44. Coplan JD, Aaronson CJ, Panthangi V, Kim Y. Treating comorbid anxiety and depression: Psychosocial and pharmacological approaches. World J Psychiatry. 2015;5:366–78.

45. Cui L, Li S, Wang S, Wu X, Liu Y, Yu W, et al. Major depressive disorder: hypothesis, mechanism, prevention and treatment. Signal Transduct Target Ther. 2024;9:30.

46. Zhao F, Cheng Z, Piao J, Cui R, Li B. Dopamine Receptors: Is It Possible to Become a Therapeutic Target for Depression? Front Pharmacol. 2022;13:947785.

47. Baldwin DS, Ajel K, Masdrakis VG, Nowak M, Rafiq R. Pregabalin for the treatment of generalized anxiety disorder: an update. Neuropsychiatr Dis Treat. 2013;9:883–92.

48. Paoletti P, Bellone C, Zhou Q. NMDA receptor subunit diversity: impact on receptor properties, synaptic plasticity and disease. Nat Rev Neurosci. 2013;14:383–400.

49. Egunlusi AO, Joubert J. NMDA receptor antagonists: Emerging insights into molecular mechanisms and clinical applications in neurological disorders. Pharmaceuticals (Basel). 2024;17:639.

50. Goldschen-Ohm MP. Benzodiazepine modulation of GABAA receptors: A mechanistic perspective. Biomolecules. 2022;12:1784.

51. Merza Mohammad TA, Merza Mohammad TA, Salman DM, Jaafar HM. Pentoxifylline as a novel add-on therapy for major depressive disorder in adult patients: A randomized, double-blind, placebo-controlled trial. Pharmacopsychiatry. 2024;57:205–14.

52. Yasrebi S-O, Momtazmanesh S, Moghaddam HS, Shahmansouri N, Mehrpooya M, Arbabi M, et al. Pentoxifylline for treatment of major depression after percutaneous coronary intervention or coronary artery bypass grafting: A randomized, double-blind, placebo-controlled trial. J Psychosom Res. 2021;150:110635.

53. Farajollahi-Moghadam M, Sanjari-Moghaddam H, Ghazizadeh Hasemi M, Sanatian Z, Talaei A, Akhondzadeh S. Efficacy and safety of pentoxifylline combination therapy in major depressive disorder: a randomized, double-blind, placebo-controlled clinical trial: a randomized, double-blind, placebo-controlled clinical trial. Int Clin Psychopharmacol. 2021;36:140–6.

54. El-Haggar SM, Eissa MA, Mostafa TM, El-Attar KS, Abdallah MS. The phosphodiesterase inhibitor pentoxifylline as a novel adjunct to antidepressants in major depressive disorder patients: A proof-of-concept, randomized, double-blind, placebo-controlled trial. Psychother Psychosom. 2018;87:331–9.

55. Maurice DH, Ke H, Ahmad F, Wang Y, Chung J, Manganiello VC. Advances in targeting cyclic nucleotide phosphodiesterases. Nat Rev Drug Discov. 2014;13:290–314.

56. Ramzi A, Maya S, Balousha N, Amin M, Powell RC, Shiha MR. Effects of the anti-inflammatory pentoxifylline on psychiatric and neuropsychiatric conditions: exploring various off-label utilities with meta-analyses. Inflammopharmacology. 2025;33:105–19.

57. Brownstein DJ, Salagre E, Köhler C. Blockade of the angio-tensin system improves mental health domain of quality of life: ameta-analysis of randomized clinical trials. Aust N Z J Psychiatry. 2018;52:24–38.

58. Vian J, Pereira C, Chavarria V, Köhler C, Stubbs B, Quevedo J, et al. The renin-angiotensin system: a possible new target for depression. BMC Med. 2017;15:144.

59. Ali NH, Al-Kuraishy HM, Al-Gareeb AI, Albuhadily AK, Hamad RS, Alexiou A, et al. Role of brain renin-angiotensin system in depression: A new perspective. CNS Neurosci Ther. 2024;30:e14525.

60. Meng X, Navoly G, Giannakopoulou O, Levey DF, Koller D, Pathak GA, et al. Multi-ancestry genome-wide association study of major depression aids locus discovery, fine mapping, gene prioritization and causal inference. Nat Genet. 2024;56:222–33.

61. Zemdegs J, Martin H, Pintana H, Bullich S, Manta S, Marqués MA, et al. Metformin promotes anxiolytic and antidepressant-like responses in insulin-resistant mice by decreasing circulating branched-chain amino acids. J Neurosci. 2019;39:5935–48.

62. Mendonça IP, Paiva IHR de,Duarte-Silva EP, Melo MG de, Silva RS da, Oliveira WH de, et al. Metformin and fluoxetine improve depressive-like behavior in a murine model of Parkinsońs disease through the modulation of neuroinflammation, neurogenesis and neuroplasticity. Int Immunopharmacol. 2022;102:108415.

63. Zhang Y, Chan VK-Y, Chan SSM, Chan EWY, Lee CH, Wong IC, et al. Effect of metformin on the risk of depression: A systematic review and meta-regression of observational studies. Asian J Psychiatr. 2024;92:103894.

64. Calkin CV, Chengappa KNR, Cairns K, Cookey J, Gannon J, Alda M, et al. Treating insulin resistance with metformin as a strategy to improve clinical outcomes in treatment-resistant bipolar depression (the TRIO-BD study): A randomized, quadruple-masked, placebo-controlled clinical trial: A randomized, quadruple-masked, placebo-controlled clinical trial. J Clin Psychiatry [Internet]. 2022;83. Available from: 10.4088/JCP.21m14022

65. Abdallah MS, Mosalam EM, Zidan A-AA, Elattar KS, Zaki SA, Ramadan AN, et al. The antidiabetic metformin as an adjunct to antidepressants in patients with major depressive disorder: A proof-of-concept, randomized, double-blind, placebo-controlled trial. Neurotherapeutics. 2020;17:1897–906.

66. Zheng P, Pan C, Zhou C, Liu B, Wang L, Duan S, et al. Contribution of Nischarin/IRAS in CNS development, injury and diseases. J Adv Res. 2023;54:43–57.

67. Smith KL, Jessop DS, Finn DP. Modulation of stress by imidazoline binding sites: implications for psychiatric disorders. Stress. 2009;12:97–114.

68. Yan S, Xu C, Yang M, Zhang H, Cheng Y, Xue Z, et al. The expression of agmatinase manipulates the affective state of rats subjected to chronic restraint stress. Neuropharmacology. 2023;229:109476.

69. Zhu MY, Wang WP, Huang J, Feng YZ, Regunathan S. Bissette Repeated immobilization stress alters rat hippocampal and prefrontal cortical morphology in parallel with endogenous agmatine and arginine decarboxylase levels Neurochem. Neurochem Int. 2008;53:346–54.

70. Zhu MY, Wang WP, Cai ZW, Regunathan S. Ordway Exogenous agmatine has neuroprotective effects against restraint-induced structural changes in the rat brain Eur. Eur J Neurosci. 2008;27:1320–32.

71. Gawali NB, Bulani VD, Gursahani MS, Deshpande PS, Kothavade PS, Juvekar AR. Agmatine attenuates chronic unpredictable mild stress-induced anxiety, depression-like behaviours and cognitive impairment by modulating nitrergic signalling pathway. Brain Res. 2017;1663:66–77.

72. Kõks S, Luuk H, Nelovkov A, Areda T, Vasar E. A screen for genes induced in the amygdaloid area during cat odor exposure. Genes Brain Behav. 2004;3:80–9.

73. Taksande BG, Kotagale NR, Tripathi SJ, Ugale RR, Chopde CT. Antidepressant like effect of selective serotonin reuptake inhibitors involve modulation of imidazoline receptors by agmatine. Neuropharmacology. 2009;57:415–24.

74. Rahangdale S, Fating R, Gajbhiye M, Kapse M, Inamdar N, Kotagale N, et al. Involvement of agmatine in antidepressant-like effect of HMG-CoA reductase inhibitors in mice. Eur J Pharmacol. 2021;892:173739.

75. Keller B, García-Sevilla JA. Dysregulation of IRAS/nischarin and other potential I1-imidazoline receptors in major depression postmortem brain: Downregulation of basal contents by antidepressant drug treatments. J Affect Disord. 2017;208:646–52.

76. Artegiani B, Lindemann D, Calegari F. Overexpression of cdk4 and cyclinD1 triggers greater expansion of neural stem cells in the adult mouse brain. J Exp Med. 2011;208:937– 48.

77. Pedraza N, Monserrat MV, Ferrezuelo F, Torres-Rosell J, Colomina N, Miguez-Cabello F, et al. Cyclin D1-Cdk4 regulates neuronal activity through phosphorylation of GABAA receptors. Cell Mol Life Sci. 2023;80:280.

78. Li C, Li X, Chen W, Yu S, Chen J, Wang H, et al. The different roles of cyclinD1-CDK4 in STP and mGluR-LTD during the postnatal development in mice hippocampus area CA1. BMC Dev Biol. 2007;7:57.

79. de Miranda AS, de Barros JLVM, Teixeira AL. Is neurotrophin-3 (NT-3): a potential therapeutic target for depression and anxiety? Expert Opin Ther Targets. 2020;24:1225–38.

80. Desole C, Gallo S, Vitacolonna A, Montarolo F, Bertolotto A, Vivien D, et al. HGF and MET: From brain development to neurological disorders. Front Cell Dev Biol. 2021;9:683609.

81. Sisi M, Fusaroli M, De Giglio A, Facchinetti F, Ardizzoni A, Raschi E, et al. Psychiatric adverse reactions to anaplastic lymphoma kinase inhibitors in non-small-cell lung cancer: Analysis of spontaneous reports submitted to the FDA adverse event reporting system. Target Oncol. 2022;17:43–51.

82. Barbieri MA, Russo G, Sorbara EE, Cicala G, Franchina T, Santarpia M, et al. Neuropsychiatric adverse drug reactions with oral tyrosine kinase inhibitors in metastatic colorectal cancer: an analysis from the FDA Adverse Event Reporting System. Front Oncol. 2023;13:1268672.

83. Xiao Z, Cao J, Wu S, Zhou T, Li C, Duan J, et al. Spectrum of psychiatric adverse reactions to cyclin-dependent kinases 4/6 inhibitors: A pharmacovigilance analysis of the FDA adverse event reporting system. CNS Neurosci Ther. 2024;30:e14862.

84. Zhu X, Nedelcovych MT, Thomas AG, Hasegawa Y, Moreno-Megui A, Coomer W, et al. JHU-083 selectively blocks glutaminase activity in brain CD11b+ cells and prevents depression-associated behaviors induced by chronic social defeat stress. Neuropsychopharmacology. 2019;44:683–94.

85. Bell BJ, Hollinger KR, Deme P, Sakamoto S, Hasegawa Y, Volsky D, et al. Glutamine antagonist JHU083 improves psychosocial behavior and sleep deficits in EcoHIV-infected mice. Brain Behav Immun Health. 2022;23:100478.

86. Liy PM, Puzi NNA, Jose S, Vidyadaran S. Nitric oxide modulation in neuroinflammation and the role of mesenchymal stem cells. Exp Biol Med (Maywood). 2021;246:2399–406.

87. Förstermann U, Sessa WC. Nitric oxide synthases: regulation and function. Eur Heart J. 2012;33:829–37, 837a – 837d.

88. Montezuma K, Biojone C, Lisboa SF, Cunha FQ, Guimarães FS, Joca SRL. Inhibition of iNOS induces antidepressant-like effects in mice: pharmacological and genetic evidence. Neuropharmacology. 2012;62:485–91.

89. Beheshti F, Hashemzehi M, Hosseini M, Marefati N, Memarpour S. Inducible nitric oxide synthase plays a role in depression- and anxiety-like behaviors chronically induced by lipopolysaccharide in rats: Evidence from inflammation and oxidative stress. Behav Brain Res. 2020;392:112720.

90. Coelho AA, Vila-Verde C, Sartim AG, Uliana DL, Braga LA, Guimarães FS, et al. Inducible nitric oxide synthase inhibition in the medial prefrontal cortex attenuates the anxiogenic-like effect of acute restraint stress via CB1 receptors. Front Psychiatry. 2022;13:923177.

91. Gill D, Georgakis MK, Walker VM, Schmidt AF, Gkatzionis A, Freitag DF, et al. Mendelian randomization for studying the effects of perturbing drug targets. Wellcome Open Res. 2021;6:16.

92. Yuan Y, Yu L, Bi C, Huang L, Su B, Nie J, et al. A new paradigm for drug discovery in the treatment of complex diseases: drug discovery and optimization. Chin Med. 2025;20:40.

93. Jia J, Zhu F, Ma X, Cao Z, Cao ZW, Li Y, et al. Mechanisms of drug combinations: interaction and network perspectives. Nat Rev Drug Discov. 2009;8:111–28.

94. Burgess S, Mason AM, Grant AJ, Slob EAW, Gkatzionis A, Zuber V, et al. Using genetic association data to guide drug discovery and development: Review of methods and applications. Am J Hum Genet. 2023;110:195–214.

95. Zhao H, Rasheed H, Nøst TH, Cho Y, Liu Y, Bhatta L, et al. Proteome-wide Mendelian randomization in global biobank meta-analysis reveals multi-ancestry drug targets for common diseases. Cell Genom. 2022;2:None.

96. Spierling SR, Zorrilla EP. Don’t stress about CRF: assessing the translational failures of CRF1antagonists. Psychopharmacology (Berl). 2017;234:1467–81.

97. Miller AH. Advancing an inflammatory subtype of major depression. Am J Psychiatry. 2025;appiajp20250289.

98. Jiang Y, Peng T, Gaur U, Silva M, Little P, Chen Z, et al. Role of corticotropin-releasing factor in the neuroimmune mechanisms of depression: Examination of current pharmaceutical and herbal therapies. Front Cell Neurosci. 2019;13:290.

