## Supplementary Materials for "Genetically informed drug target prioritisation and repurposing for major depressive disorder"

### Supplementary Methods

#### *GWAS summary statistics*

All analyses were performed using the human reference genome GRCh38 reference assembly, and GWAS data were harmonised using the Python package *gwas-norm* [1]. For proteomic data, where possible, we selected GWAS of protein values measured using different assay platforms. Due to limited data availability in the brain and CSF, this was only feasible for blood plasma. We selected the two most well-powered blood plasma GWAS of protein values measured using two commonly used high-throughput proteomic platforms (deCODE using SomaScan and UK Biobank Pharma Proteomics Project [UKB-PPP] using Olink) [2,3]. While these platforms show only modest correlations in their protein measurements due to different binding mechanisms and protein measure normalisation approaches, using both provides complementary information [4]. First, it enables testing non-overlapping proteins unique to each platform, improving overall protein coverage. Second, for overlapping proteins measured on both platforms, concordant findings provide stronger evidence of reliable protein quantification [4]. We deprioritised blood pQTL targets that showed significant but directionally opposite MR-IVW/-Wald effect estimates between our deCODE and UKB-PPP. These panels measure the same molecular trait (proteins) in the same tissue type (blood plasma) but using different protein assay platforms. Thus, finding opposite directions between these panels could indicate potential technical artefacts in protein measurement. We did not penalise targets for directional inconsistencies between other tissues or molecular trait modalities, as such differences might reflect genuine biological variation across tissues or in protein versus gene expression rather than technical artefacts.

##### *Drug target selection and druggability assessment*

We used Open Targets tractability data to define the druggable genome (version dated 2024-05-23). This framework assessed the druggable potential (or 'druggability') of 18,868 protein-coding genes based on their amenability to four drug modalities. This included small molecules, monoclonal antibodies [5], an emerging therapeutic approach called proteolysis targeting chimeras (PROTACs) [6], and 'other modalities' (e.g. biologics, oligonucleotides). Open Targets incorporated clinical trial evidence from ChEMBL for all four modality groups. ChEMBL targets of drugs and/or clinical trial compounds encompassed both single proteins or multiple proteins functioning as a protein complex or protein family [7]. For experimental druggability assessments, this included data for small molecules, antibodies, and PROTACs. Experimental data were drawn from various databases incorporated by Open Targets, including UniProt, Human Protein Atlas, Protein Data Bank, and DrugEBIlity, evaluating factors such as protein structure, cellular location, and previous research evidence (see further details below). Our approach yielded 3,652 genes encoding drug target proteins, comprising 1,452 targets with clinical trial evidence (935 phase IV, 489 phase II-III, 28 phase I) and 2,200 with high-quality experimental evidence.

We retained targets meeting at least one tractability assessment across the four modalities, each with hierarchical bucket scores. The 'clinical precedence' category comprised bucket 1 (targets of phase IV drugs), bucket 2 (targets of compounds in advanced clinical trials, phases II-III), and bucket 3 (targets of compounds in early clinical trials, phase I). For experimental evidence, we included targets with the highest quality assessments (bucket 4) within each modality. Targets with experimental evidence for small-molecule binding required the highest 'discovery precedence' category evidence score of 1. This included targets with both structural ligand evidence (co-crystallised structure with a small molecule) and compound quality evidence (high-quality ligands meeting drug-like criteria). Targets with evidence for antibody-based therapy required a 'predicted tractable high confidence evidence' score of 1, with a high confidence accessible subcellular location from both UniProt and Gene Ontology classifications. PROTAC targets required literature precedence (documented in PROTAC development publications) combined with high confidence locations for the PROTAC approach (cytoplasm/nucleus location [scores 1-2]). Details on tractability bucket criteria are available in the Open Targets tractability pipeline documentation at https://github.com/chembl/tractability_pipeline_v2 and https://platform-docs.opentargets.org/target/tractability.

#### *MR-IVW robustness*

For generalised least squares MR-Egger, which allows for correlated instruments, we applied an LD R-squared threshold of 0.4 using the same random sample of 5,000 UKB participants as our primary analysis to generate correlation matrices. For contamination mixture and weighted median models, we used a more stringent clumping threshold of 0.01. We restricted robust MR estimates to targets with at least 3 IVs, as this is the minimum number of IVs required for both MR-Egger and weighted median methods. For MR-Egger, we used a random-effects model [8].

#### *External pQTL replication*

We used the same parameters for replication analyses as those applied in our primary *cis*-MR analysis. For blood pQTL targets significant in our discovery panels (deCODE and UK Biobank), we tested for replication in two independent blood pQTL panels (INTERVAL and Gudjonsson) [9,10]. Both replication panels use SomaScan platforms, providing within-platform replication for deCODE results, while also providing cross-platform replication for the UKB-PPP Olink measurements. Brain pQTL targets identified in ROSMAP were tested for replication in the Banner pQTL panel, as these panels match in protein assay platform and brain regions [11]. We note there was no overlap between targets identified in the discovery blood pQTL and brain pQTL panels. We therefore applied tissue-specific Bonferroni corrections, with separate thresholds for blood and brain based on the number of targets tested in replication panels. For blood pQTLs, we accounted for targets tested more than once in blood replication panels by correcting for the total number of tests (Supplementary Table 3).

#### *Colocalisation*

We set the Bayesian prior probability that a variant is associated with either the molecular trait (p1) or MDD (p2) individually as $p1 = p2 = 1\times10^{-4}$ and a lower prior probability for shared associations with both traits ($p12 = 1\times10^{-5}$), consistent with the recommended threshold for eQTL studies [12]. We set the genomic window size to match that of our *cis*-

MR analyses [13]. For each target, we assessed the statistical power of our colocalisation analysis by examining the sum of posterior probabilities for hypotheses 3 and 4 (PPH3 + PPH4). A low combined probability for these hypotheses indicates limited statistical power to detect colocalisation [14,15]. We therefore established a power threshold where the sum of PPH3 and PPH4 needed to reach at least 0.8 to consider the analysis sufficiently powered [15]. This threshold helped us distinguish between true negative results and those arising from insufficient statistical power.

##### *Estimated therapeutic relevance*

We estimated the therapeutic relevance of each compound for MDD by integrating drug mechanisms of action data from ChEMBL [7] with our primary MR results using the *bio-misc* package. We classified drug mechanisms of action as either activation or inhibition based on their mechanism of action on target proteins. Activation mechanisms included agonists, partial agonists, activators, positive allosteric modulators, and openers. Inhibition mechanisms comprised antagonists, allosteric antagonists, blockers, inverse agonists, negative allosteric modulators, antisense inhibitors, degraders, and inhibitors.

#### *Known depression-related drug effects*

Depression-related drug effects could indicate: (1) for primary therapeutic indications: the compound paired to the target is relevant for the treatment of depression-related symptoms, ranging from early-stage clinical trials to approved medications (phases I-IV); (2) for side effects, cautions, or contraindications: depression-related effects are reported for the compound. We developed two distinct sets of terms to classify drug effects: core depression terms and terms more broadly related to depression based on diagnostic criteria. Core depression terms captured primary depression-associated drug effects, including major depressive disorder, depression, apathy, mood, anxiety, and suicidality. The related depression terms encompass broader psychiatric and somatic symptoms associated with depression. This included changes in activity, appetite, social behaviour, concentration, weight, restlessness, and sexual interest (i.e. loss of interest or pleasure). Broader but related terms that captured the effects of psychiatric medication included ‘psychiatric’, ‘abnormal thinking’, ‘negative symptoms’ (of psychosis and schizophrenia that overlap with depression) and ‘feeling abnormal’.

*Positive control analysis*

We were able to test in *cis*-MR targets for 61 approved compounds and 169 clinical trial compounds. Compounds approved for one indication (e.g. MDD) but in clinical trials for another (e.g. anxiety) were classified only in the approved category. At the target level, we tested 80 approved PC targets and 205 clinical PC targets. We note that 76 approved PC targets overlapped with clinical trial PC targets, reflecting that targets of approved drugs are also targets of other compounds tested in clinical trial phases I-III.

We constructed contingency tables for brain and blood tissues separately, counting targets with significant and non-significant *cis*-MR estimates in each category. Comparisons were restricted to brain and blood due to the limited number of targets found in CSF. Odds ratios (OR) quantified enrichment, calculated as: (PC target significant/other target significant)/(PC non-significant target/other non-significant target). Statistical significance was tested using chi-square tests, with Yates' correction applied when expected cell counts were < 5, and a p-value < 0.05 considered statistically significant.

#### References

1. Schmidt AF, Bourfiss M, Alasiri A, Puyol-Anton E, Chopade S, van Vugt M, et al. Druggable proteins influencing cardiac structure and function: Implications for heart failure therapies and cancer cardiotoxicity. Sci Adv. 2023;9:eadd4984.

2. Ferkingstad E, Sulem P, Atlason BA, Sveinbjornsson G, Magnusson MI, Styrmisdottir EL, et al. Large-scale integration of the plasma proteome with genetics and disease. Nat Genet. 2021;53:1712–21.

3. Sun BB, Chiou J, Traylor M, Benner C, Hsu Y-H, Richardson TG, et al. Plasma proteomic associations with genetics and health in the UK Biobank. Nature. 2023;622:329–38.

4. Eldjarn GH, Ferkingstad E, Lund SH, Helgason H, Magnusson OT, Gunnarsdottir K, et al. Large-scale plasma proteomics comparisons through genetics and disease associations. Nature. 2023;622:348–58.

5. Brown KK, Hann MM, Lakdawala AS, Santos R, Thomas PJ, Todd K. Approaches to target tractability assessment - a practical perspective. Medchemcomm. 2018;9:606–13.

6. Schneider M, Radoux CJ, Hercules A, Ochoa D, Dunham I, Zalmas L-P, et al. The PROTACtable genome. Nat Rev Drug Discov. 2021;20:789–97.

7. Mendez D, Gaulton A, Bento AP, Chambers J, De Veij M, Félix E, et al. ChEMBL: towards direct deposition of bioassay data. Nucleic Acids Res. 2019;47:D930–40.

8. Patel A, Ye T, Xue H, Lin Z, Xu S, Woolf B, et al. MendelianRandomization v0.9.0: updates to an R package for performing Mendelian randomization analyses using summarized data. Wellcome Open Res. 2023;8:449.

9. Gudjonsson A, Gudmundsdottir V, Axelsson GT, Gudmundsson EF, Jonsson BG, Launer LJ, et al. A genome-wide association study of serum proteins reveals shared loci with common diseases. Nat Commun. 2022;13:480.

10. Sun BB, Maranville JC, Peters JE, Stacey D, Staley JR, Blackshaw J, et al. Genomic atlas of the human plasma proteome. Nature. 2018;558:73–9.

11. Robins C, Liu Y, Fan W, Duong DM, Meigs J, Harerimana NV, et al. Genetic control of the human brain proteome. Am J Hum Genet. 2021;108:400–10.

12. Wallace C. Eliciting priors and relaxing the single causal variant assumption in colocalisation analyses. PLoS Genet. 2020;16:e1008720.

13. Hukerikar N, Hingorani AD, Asselbergs FW, Finan C, Schmidt AF. Prioritising genetic findings for drug target identification and validation. Atherosclerosis. 2024;390:117462.

14. Giambartolomei C, Vukcevic D, Schadt EE, Franke L, Hingorani AD, Wallace C, et al. Bayesian test for colocalisation between pairs of genetic association studies using summary statistics. PLoS Genet. 2014;10:e1004383.

15. Guo H, Fortune MD, Burren OS, Schofield E, Todd JA, Wallace C. Integration of disease association and eQTL data using a Bayesian colocalisation approach highlights six candidate causal genes in immune-mediated diseases. Hum Mol Genet. 2015;24:3305–13.
